# Distinct gut microbiome shifts in the NICU influence later atopic dermatitis development

**DOI:** 10.1101/2025.09.03.25334680

**Authors:** Emily S Robbins, Kathryn E McCauley, Rachel Strength, Sivaranjani Namasivyam, Angelina G Angelova, Ruhika Prasad, Anal Patel, Varsha Deopujari, Andrew S Burns, Shreni Mistry, Rajiv Baveja, Robin L Baker, Pamela A Frischmeyer-Guerrerio, Shira Levy, Suchitra K Hourigan

**Author notes:** Corresponding author: Suchitra Hourigan. Laboratory of Host Immunity and Microbiome, National Institute of Allergy and Infectious Diseases, National Institutes of Health, Building 50, Rm 5511, 50 South Dr, Bethesda, MD, USA, 20892.

## Abstract

While it has previously been shown that early life gut microbiome imbalance is associated with the later development of atopic dermatitis (AD) in full term infants, to our knowledge no similar studies have been conducted in infants admitted to the Neonatal Intensive Care Unit (NICU). This is an important research gap to address because a) infants in the NICU are at an increased risk for gut microbiome dysbiosis due to several clinical and environmental factors including frequent antimicrobial exposure; and b) their risk of developing AD may be different from full term infants. We demonstrate gut microbiome dysbiosis and clinical factors impacting the microbiome associated with later development of AD that are unique to infants in the NICU. These may represent early predictors of AD development and potential therapeutic targets to mitigate future disease.

## MAIN

It has previously been shown that early life gut microbiome dysbiosis is associated with the later development of atopic dermatitis (AD) in full term healthy infants^1^. However, while early life skin microbiome changes have been associated with the later development of AD in infants admitted to the neonatal intensive care unit (NICU), to our knowledge no similar studies have been conducted examining the gut microbiome of infants in the NICU^2^. This is an important research gap to address because a) infants in the NICU are at an increased risk for gut microbiome dysbiosis due to several clinical and environmental factors including frequent antimicrobial exposure; and b) their risk of developing AD may be different from full term infants^3,4^.

To address this, we conducted shotgun metagenomic sequencing of 877 serial stool samples collected over 3 years from 128 infants admitted to the NICU from an ongoing longitudinal cohort study^2,5^. Of the 113 infants that had AD outcome data, 40 (35%) developed AD by the age of 5 (additional details in Supplementary Methods). Infants with AD were more likely to have received antibiotics while they were in the NICU (p=0.017) (full demographic and clinical details of infants who develop AD versus no AD in Supplementary Table 1). While no differences in alpha diversity were seen between those with and without AD over time (Figure 1A), which differs from some studies in full term infants^6^, the microbiome composition between the two groups was significantly different in early life, with those who later developed AD shifting away from no AD (Figure 1B). The two groups converged in composition around day of life 150. Unsupervised longitudinal clustering with latent class mixed models identified trajectories of microbiota development, and the second principal coordinate divided into two independent trajectories (Figure 1C, additional details in Supplementary Methods), of which trajectory B was enriched in *Escherichia* and associated with an increased risk of developing AD [OR 3.04 (1.05, 9.19), p=0.043], adjusted for NICU length of stay, delivery mode, maternal asthma, infant sex and gestational age, continuing to trend after adjustment for antibiotic administration [OR 2.56 (0.861, 7.90), p=0.0935]. At a species level, several *Enterobacteriaceae* species were found in the earliest days of life in infants who developed AD (Figure 1D, E). Some of these species, including *Escherichia spp*, are found in full term infants who develop AD^1^, but with notable differences including lack of a *Klebsiella* association with AD in infants in the NICU, despite this organism being highly abundant in samples from this NICU cohort (median abundance = 113,339 reads per million; dominant genus in 15% of samples). Assessment of bacterial pathways annotated by KEGG Orthology identified several significant pathways to be associated with AD (Figure 1F, G), most notably from the flagellar assembly pathway, with bacterial flagellin previously identified as promoting atopic disease^7^.

**Figure 1.**
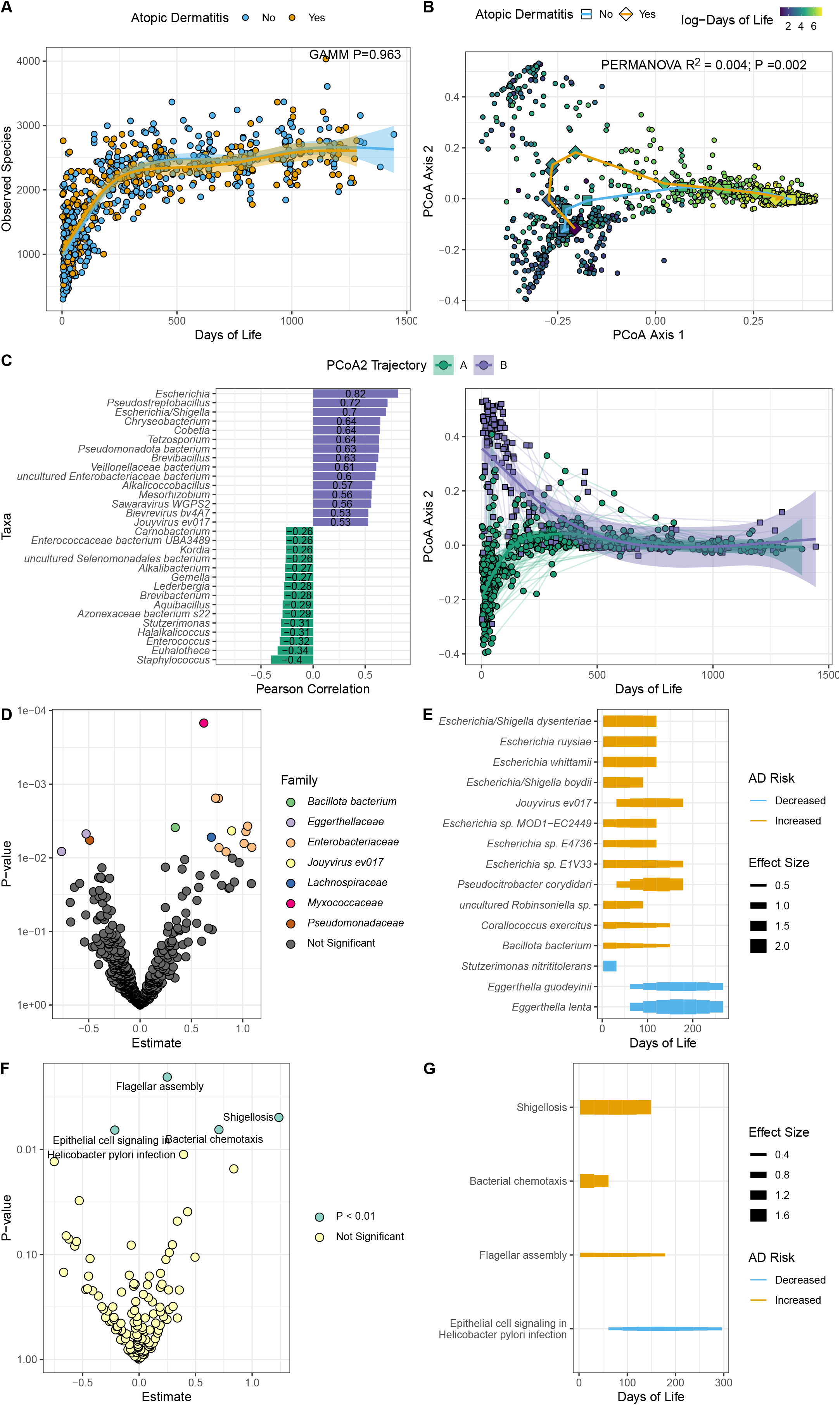
(A) Observed Species over Days of Life (B) Sample Bray Curtis distance with longitudinal change as deciles of Days of Life for no AD (circles) and AD (squares) (C) Pearson correlation of species with PCoA Axis 2; PCoA Axis 2 over time, showing LCMM groups (A green, B purple). (D) Differential species by AD using GAMM; points are colored by bacterial family. (E) Effect size over time among significant taxa (F) Differential pathways by AD using GAMM. (G) Effect size over time among significant pathways. Abbreviations: AD: Atopic Dermatitis; GAMM: Generalized Additive Mixed Models; LCMM: Latent Class Mixed Models; PCoA: Principal Coordinates Analysis

We next examined clinical factors that are mediated by trajectory B in their AD risk. Vaginal delivery, chorioamnionitis and preterm labor exhibited an indirect relationship with AD through trajectory B (Figure 2). In contrast, several but not all studies have shown an increased risk of AD with Cesarean Section delivery in full term infants, with the hypothesis that the infant exposure to the maternal vaginal/gut microbiome in a vaginal delivery may be protective^8^. It is possible that the vaginal microbiome dysbiosis known to be associated with a premature birth may not provide the same protection^9^. Chorioamnionitis and preterm labor may also represent factors more common in infants in the NICU, and not previously seen in full term studies. Of note, unlike full term studies, breast milk use is almost universal in the NICU and so the contribution of this factor is difficult to assess.

**Figure 2.**
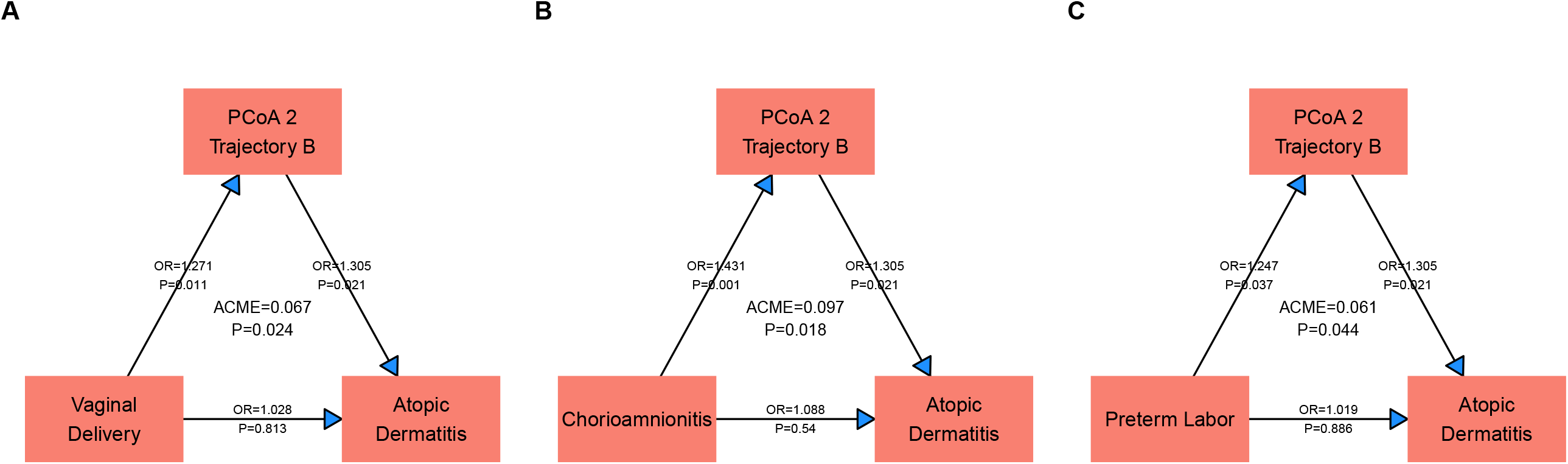
Directed Acyclic Graphs for factors in which the PCoA 2 Trajectory served as mediator to Atopic Dermatitis, including (A) Vaginal Delivery (B) Chorioamnionitis and (C) Preterm Labor. Abbreviations: ACME: Average Causal Mediation Effect, OR: Odds Ratio, PCoA: Principal Coordinates Analysis

In conclusion, this study demonstrates gut microbiome changes and clinical factors impacting the microbiome associated with AD that are unique to infants in the NICU. These may represent early predictors of AD development and potential therapeutic targets to mitigate further disease.

## Data Availability

All data produced in the present study are available upon reasonable request to the authors

https://dataview.ncbi.nlm.nih.gov/object/PRJNA1280936?reviewer=uvl549kes57nrjq1pco2h6hklc

## Supplementary Data

Supplementary Table 1: Comparison of maternal and infant characteristics between atopic dermatitis (AD) and non-atopic dermatitis (Non-AD) groups

Supplementary Methods

## Acknowledgments

The content is solely the responsibility of the authors and does not necessarily represent the official views of the National Institutes of Health. The authors thank Kristy and Roger Crombie for their generous philanthropic donation toward this project in loving memory of their daughter Anna Charlotte. The authors thank the NIH Intramural Sequencing Center (NISC) for assistance with sequencing. This study used the Office of Cyber Infrastructure and Computational Biology (OCICB) High Performance Computing (HPC) cluster at the National Institute of Allergy and Infectious Diseases (NIAID), Bethesda, MD.

## Ethics approval and consent to participate

All subjects provided signed informed consent prior to collection and storage of samples. This study was Institutional Review Board approved (WCG IRB 1300205). Consent forms included language authorizing publication of findings.

## Author Contributions

Conceptualization: ER, RBav, RBak, PAG, SL, SKH

Methodology: ER, KM, RS, SN, SL, SKH

Investigation: ER, KM, RS, SN, AA, RP, AP, VD, ASB, SM, RBav, RBak, PAG, SL, SKH

Data Curation: ER, KM, RS, SN, RP, SL, SKH

Visualization: ER, KM, SN

Funding acquisition: SKH

Project administration: SL, SKH

Supervision: SKH

Writing – original draft: ER, KM, SKH

Writing – review & editing: ER, KM, RS, SN, AA, RP, AP, VD, ASB, SM, RBav, RBak, PAG, SL, SKH

## Data and code availability

Sequencing reads are deposited in the Sequence Read Archive (SRA) under submission PRJNA1280936.

This paper does not report original code.

