## Supplemental Table 1 for "Distinct gut microbiome shifts in the NICU influence later atopic dermatitis development"

Supplementary Table 1: Comparison of maternal and infant characteristics between atopic dermatitis (AD) and non-atopic dermatitis (Non-AD) groups

^#^p-values are based on Fisher or Chi-square tests for categorical variables, unpaired t-test for normally distributed continuous variables and Mann Whitney test for non-normally distributed data

| **Maternal Characteristics** | | | |
| --- | --- | --- | --- |
|  | **AD (n=40)** | **Non-AD (n=73)** | **p-value^#^** |
| **Maternal Age in years, mean (±SD)** | 34.8 (±5.57) | 33.5 (±5.42) | 0.2426 |
| **Maternal Ethnicity** | | | |
| Hispanic or Latino | 4 (10%) | 6 (8%) | 0.7516 |
| Non-Hispanic or Latino | 30 (75%) | 52 (71%) |  |
| Declined to answer/Unknown | 6 (15%) | 15 (21%) |  |
| **Maternal Race** | | | |
| Caucasian | 23 (57.5%) | 38 (52%) | 0.5689 |
| Asian | 5 (12.5%) | 15 (21%) |  |
| African American or Black | 4 (10%) | 10 (14%) |  |
| More than one, Other, decline to answer/unknown | 8 (20%) | 10 (14%) |  |
| **Mode of Delivery** | | | |
| Vaginal | 8 (20%) | 13 (18%) | 0.8037 |
| Cesarean Section | 32 (80%) | 60 (82%) |  |
| **Labor** | | | |
| Pre-term | 6 (15%) | 10 (14%) | >0.9999 |
| Not pre-term | 34 (85%) | 63 (86%) |  |
| **Peripartum Antibiotics** | | | |
| Yes | 38 (95%) | 69 (95%) | >0.9999 |
| No | 2 (5%) | 4 (5%) |  |
| **Chorioamnionitis** | | | |
| Yes | 6 (15%) | 8 (11%) | 0.56 |
| No | 34 (85%) | 65 (89%) |  |
| **Maternal Asthma or Inhaler** |  |  |  |
| Yes | 4 (10%) | 9 (12%) | >0.9999 |
| No | 36 (90%) | 64 (88%) |  |
| **Maternal Smoking** |  |  |  |
| Yes, before pregnancy | 7 (17.5%) | 12 (16%) | 0.7279 |
| Never | 30 (75%) | 52 (71%) |  |
| Unknown | 3 (7.5%) | 9 (12%) |  |
| **Infant Characteristics** | | | |
|  | **AD (n=40)** | **Non-AD (n=73)** | **p-value^#^** |
| **Gestational Age** |  |  |  |
| Extremely Preterm (<28 weeks) | 8 (20%) | 10 (14%) | **0.0391** |
| Very Preterm (28-<32 weeks) | 9 (22.5%) | 24 (33%) |  |
| Moderate Preterm (32-<34 weeks) | 4 (10%) | 20 (27%) |  |
| Late Preterm (34-<37 weeks) | 12 (30%) | 15 (21%) |  |
| Term (≥37 weeks) | 7 (17.5%) | 4 (5%) |  |
| **Infant Sex** |  |  |  |
| Male | 15 (13%) | 34 (30%) | 0.3311 |
| Female | 25 (22%) | 39 (35%) |  |
| **Length of Stay in NICU in days, mean (±SD)** | 50.7 (±40.77) | 47.5 (±40.15) | 0.606 |
| **NICU Antibiotics** | | | |
| Yes | 29 (72.5%) | 35 (47.9%) | **0.0168** |
| No | 11 (27.5%) | 38 (52.1%) |  |
| **Antibiotics in the first 30 days of life** | | | |
| Ampicillin and Gentamicin only | 20 (50%) | 25 (34.2%) | 0.3096 |
| Ampicillin, Gentamicin and Other | 8 (20%) | 7 (9.6%) |  |
| Others | 0 (0%) | 3 (4.1%) |  |
| **Post Discharge Antibiotics** | | | |
| Yes | 6 (15%) | 8 (11%) | 0.3824 |
| No | 34 (85%) | 61 (84%) |  |
| Unknown | 0 (0%) | 4 (5%) |  |
| **Breastmilk in NICU** | | | |
| Yes | 29 (72.5%) | 60 (82%) | 0.3296 |
| No | 1 (2.5%) | 2 (3%) |  |
| Unknown | 10 (25%) | 11 (15%) |  |
| **Breastmilk Post Discharge** | | | |
| Yes | 18 (45%) | 25 (34%) | 0.2839 |
| No | 22 (55%) | 44 (60%) |  |
| Unknown | 0 (0%) | 4 (5%) |  |
| **Siblings at home?** | | | |
| none | 13 (32.5%) | 19 (26%) | 0.614 |
| one | 16 (40%) | 26 (36%) |  |
| two | 8 (20%) | 15 (21%) |  |
| three or more | 3 (7.5%) | 9 (12%) |  |
| unknown | 0 (0%) | 4 (5%) |  |
| **Cat at home?** | | | |
| Yes | 3 (7.5%) | 2 (3%) | 0.3705 |
| No | 15 (37.5%) | 23 (32%) |  |
| Unknown | 22 (55%) | 48 (66%) |  |
