## Supplemental Methods for "Distinct gut microbiome shifts in the NICU influence later atopic dermatitis development"

*Subjects and Sample Collection*

Neonates who spent time in the Inova Fairfax Neonatal Intensive Care Unit (NICU) were enrolled in an observational longitudinal microbiome cohort study, as previously described^1,2^. The study was Institutional Review Board approved (WCG IRB 1300205) and parental informed consent was obtained. Neonates were enrolled within the first week of life and had an anticipated stay in the NICU of >5 days. While in the NICU stool samples were collected for microbiome analysis up to twice a week, and stored as whole stool at -80°C until analysis. Detailed demographic and clinical data was collected including delivery mode, gestational age, maternal peripartum antibiotic use and infant antibiotics.

After discharge from the NICU, follow-up surveys reporting health, illnesses, diet and a variety of exposures were collected approximately every 3–6 months until approximately 3 years of age (corrected for gestational age), accompanied by a stool sample collected by previously validated methods^3^. One final follow up survey was sent to every family when the child was around age 5 years of age, with more specifics regarding atopic diagnoses. For this, a slightly modified ISAAC questionnaire was used to determine the diagnosis of atopic dermatitis (AD), allergic rhinitis, asthma, or food allergies^4^.

For this current study, neonates from the larger cohort were included if they had at least 1 stool sample from before 1 month of life, at least 2 stool samples overall and completed the final survey regarding allergic disease.

*DNA Extraction*

DNA was extracted from fecal samples in two stages. First, approximately 50 mg of fecal material and 650 μL MBL lysis buffer from the PowerMicrobiome DNA/RNA EP Kit (Qiagen) were added to Lysis Matrix E (LME) tubes (MP Biomedicals). LME tubes were transferred to a Precelleys 24 Tissue Homogenizer (Bertin Technologies) and fecal samples were homogenized, centrifuged, with the resultant supernatant transferred to a deep-well 96-well plate. The second stage consisted of DNA isolation from the above supernatant using the MagAttract PowerMicrobiome DNA/RNA EP Kit (Qiagen) on an automated liquid handling system as detailed by the manufacturer (Eppendorf).

*Shotgun Metagenomic Sequencing*

Total gene content of the microbiome was assessed through shotgun metagenomic sequencing. Metagenomic libraries were constructed from 100 ng of DNA as starting material using the Illumina DNA Prep kit. Illumina DNA/RNA UD Indexes were used to add sample-specific sequencing indices to both ends of the libraries. An Agilent 4200 TapeStation system with High Sensitivity D5000 ScreenTape (Agilent Technologies, Inc) was used to verify quality and assess final library size. A positive control (MSA-2002 20 Strain Even Mix Whole Cell Material (ATCC)) and a buffer extraction negative control were included. Metagenomic libraries were normalized and pooled at an equimolar concentration. Final pools were sequenced on a NovaSeq X sequencer using a paired-end (150x150) NovaSeq 25B flow cell across ten lanes (Illumina, Inc).

*Sequence Processing*

*Quality Assessment*. Paired-end sequences were assessed for quality with FastQC and MultiQC^5,6^. Reads then underwent the Whole-Genome Sequence Assembly 2 (WGSA2) protocol from the Nephele platform^7,8^. In brief, reads were processed with fastp and minimal trimming and filtering by ensuring an average read quality of 10, a trim of the 3’ end of the read at a quality of 15 and trimming the 5’ end at a Q score of 20 with additional filtering of reads if they were less than 60bp after trimming, and automatic trimming of adapters^9^. Human reads were decontaminated from sequence data using Kraken2 with a database containing the human and mouse genome^10^.

*Assembly and Gene Annotation*.

Within the WGSA2 pipeline, reads were assembled into contiguous sequences, or contigs, using metaSPAdes.^11^ Reads were recruited back to contigs using bowtie2 and SAMtools to produce information on scaffold coverage and quality^12,13^. Protein coding regions (CDS) were predicted from assembled scaffolds using Prodigal^14^. Predicted CDS regions were processed by EggNOG-mapper2 to identify and annotate genes with KEGG Orthology (KO) identifiers^15,16^. Annotated genes were agglomerated into pathways using MinPath. Non-microbial pathways were filtered by identifying all pathways present across all bacteria, fungi and archaea in the KEGG database. Abundances were calculated using VERSE to obtain Transcripts per Million (TPM) at the CDS level and summed to obtain TPM by pathway^17,18^.

*Taxonomic Classification*.

Processed reads were classified taxonomically using Ganon^19^. Three custom, independent databases were built on December 19, 2024 including (1) archaea, bacteria and fungi reference genomes from RefSeq, (2) Viral Complete Genomes from RefSeq and (3) the top 1 species of archaea and bacteria from the Genome Taxonomy DataBase (GTDB). Databases were used in a hierarchical manner, in which both RefSeq databases were used as one database, and if a read could not be classified by RefSeq, it would undergo classification by GTDB. A relative cut-off of 0.2 was used for all samples. After classification by RefSeq and GTDB, the MultiTax package, included with the Ganon package, standardized the calls to the NCBI lineage.

*Microbiome Analysis*

Taxonomy. Ganon classification resulted in 59,191 species across all samples, with samples containing between 29M and 39M (IQR) reads per sample. Species were filtered if they had fewer than 10,000 reads, resulting in 5,895 species, but only removing 0.21% of the total reads in the dataset (remaining Q1: 29M and Q3: 38M). Within samples, a minimum of 0.02% and a maximum of 4.73% of reads were removed (Q1: 0.12%; Q3: 0.25%). Samples underwent rarefaction curves and found that most samples approached an asymptote and thus abundances were normalized to reads per million to maintain all samples. Alpha diversity statistics and beta diversity distance matrices were calculated by the phyloseq package^20^.

Latent Classes. Infants were separated into distinct trajectories of microbiota development, as described by Bray Curtis PCoA 2, utilizing latent class mixed models with the lcmm package in R^21^. The 10 most dominant genera, as well as PCoA 1 and PCoA 2 from Bray Curtis were tested for the presence of divergent trajectories, though only PCoA 2 provided well-distributed, distinct trajectories that also related to AD development. In brief, a model with no separation was built to obtain initial values. Between 2 and 5 separations were performed and fit was compared to the initial values using the Bayesian Information Criterion (BIC) and log-likelihood.

Mediation Analysis. Mediation effects between all variables collected from participants at the subject level (“treatment”) and PCoA Trajectory (“Mediator”) on Atopic Dermatitis (“outcome”) were identified using the mediation package in R^22^. Variables were not considered at all if they had a prevalence less than 10%, or were missing in at least 60% of participants. Several estimates were obtained, including (1) the effect of exposure on the mediator, (2) the effect of exposure on outcome, adjusted for the mediator, (3) effect of treatment on the outcome, and (4) effect of mediator on the outcome. Each effect was estimated with generalized linear models, with additional adjustment for sex. Models 1 and 2 were used in the mediate function to calculate Average Causal Mediation Effects (ACME) and ACME p-values.

Statistical Analysis. Because the microbiota changed significantly over time due to the well-described nature of microbiome development, linear models could not adequately capture the change in features of interest over time. Thus, Generalized Additive Mixed Models from the mgcv package were used for statistical tests occurring over time in order to capture the variation specific to our variables of interest^23^. The model was constructed as follows:
Abundance ~ Variable + s(Days of Life, by= Variable) + s(SubjectID, bs=”re”)

With smoothing of days of life stratified by our variable of interest, and an additional smooth term for subject with a random effect basis. P-values less than 0.01 were considered significant, and significant intervals were calculated with the *marginal effects* package^24^; intervals were corrected for multiple comparisons and an FDR p-value < 0.05 considered significant. Permutational Analysis of Variance was calculated with the adonis2 function in vegan using an interaction term between the variable of interest and Days of Life^25^. Permutations were constrained within subjects using a “series” permutation design.
